# Mosaic loss of chromosome Y characterizes late-onset rheumatoid arthritis and contrasting associations of polygenic risk score based on age at onset

**DOI:** 10.1101/2024.09.12.24313215

**Authors:** Shunsuke Uchiyama, Yuki Ishikawa, Katsunori Ikari, Suguru Honda, Keiko Hikino, Eiichi Tanaka, Yoichiro Kamatani, Takahisa Gono, Giulio Genovese, Masataka Kuwana, Chikashi Terao

**Affiliations:** Department of Allergy and Rheumatology, Nippon Medical School Graduate School of Medicine, Tokyo, Japan; Laboratory for Statistical and Translational Genetics, RIKEN Center for Integrative Medical Sciences, Yokohama, Japan; Department of Orthopedic Surgery, Tokyo Women’s Medical University School of Medicine, Tokyo, Japan; Division of Rheumatology, Department of Internal Medicine, Tokyo Women’s Medical University School of Medicine, Tokyo, Japan; Laboratory for Pharmacogenomics, RIKEN Center for Integrative Medical Sciences, Yokohama, Japan; Department of Computational Biology and Medical Sciences, Graduate School of Frontier Sciences, The University of Tokyo, Japan; Program in Medical and Population Genetics, Broad Institute of MIT and Harvard, Cambridge, MA, USA; Department of Genetics, Harvard Medical School, Boston, MA, USA; Clinical Research Center, Shizuoka General Hospital, Shizuoka, Japan

**Author notes:** Correspondence author: Masataka Kuwana, MD, PhD Department of Allergy and Rheumatology, Nippon Medical School Graduate School of Medicine, 1-1-5 Sendagi, Bunkyo-ku, Tokyo, 113-8603, Japan, Chikashi Terao MD, PhD Laboratory for Statistical and Translational Genetics, Center for Integrative Medical Sciences, Riken, Yokohama 230-0045, Japan.

**Keywords:** mosaicism, aging, elderly, rheumatoid arthritis

## Abstract

**Objectives:** mosaic chromosomal alterations (mCAs) increase with age and are associated with age-related diseases. The association between mCAs and rheumatoid arthritis (RA), particularly late-onset rheumatoid arthritis (LORA), has not been explored.

**Methods:** mCAs were detected in peripheral blood samples from two independent Japanese datasets (Set 1:2,107 RA cases and 86,998 controls; Set 2:2,359 RA cases and 86,998 controls). The associations between mCAs and RA were evaluated in each dataset using logistic regression models and meta-analysis. In each dataset, we evaluated the effect sizes of mosaic loss of Y (mLOY) and polygenic risk score (PRS) of RA in males and subsequently performed a meta-analysis. The interaction between mLOY and PRS was assessed. These models were applied separately to RA, LORA, and young-onset RA (YORA).

**Results:** mLOY increased significantly in LORA (OR=1.43, P=0.0070). We observed a negative association between mLOY and YORA (OR=0.66, P=0.0034). On the other hand, we found consistently negative associations of autosomal mCAs or mosaic loss of X (mLOX) with RA, LORA, and YORA. The effect sizes of PRS were lower for LORA than for YORA. mLOY with a high cell fraction strengthened the association between PRS and LORA (P=0.0036), whereas the association with YORA was independent of mLOY.

**Conclusion:** LORA was characterised by the presence of a high burden of mLOY. The observed interaction between mLOY and PRS in LORA, but not in YORA, supports different gene-environment interactions between the subsets. These data suggest that distinct pathophysiological mechanisms underlie the development of LORA and YORA.

**Key messages:** *What is already known about this subject?:*

□ The prevalence of late-onset rheumatoid arthritis (LORA) in aging societies is increasing worldwide.
□ LORA has unique clinical characteristics, including higher prevalence in males, acute onset of the disease, and lower proportion of seropositivity compared with young-onset RA (YORA), suggesting distinct genetic and environmental components between the RA subsets.
□ Mosaic chromosomal alterations (mCAs) accumulate with age and are associated with age-related disorders. mCAs occur in both autosomes and sex chromosomes and would reflect the clonal expansion of dysregulated immune cells.

*What does this study add?:*

□ Mosaic loss of Y (mLOY) is increased in male patients with LORA.
□ While mLOY is positively associated with LORA, mLOY is negatively associated with YORA.
□ The association of the RA-polygenic risk score (PRS) with LORA was more apparent in the presence of mLOY, while the association with YORA was independent of mLOY.

*How might this impact clinical practice or future developments?:*

□ mLOY plays an acquired risk for RA in males by modifying the contribution of the genetic risk to RA development depending on age at onset.
□ Further investigation of the different effects of mLOY on LORA and YORA would lead to a better understanding of the pathological basis of RA influenced by aging and sex.

## INTRODUCTION

Rheumatoid arthritis (RA) is the most common form of inflammatory arthritis in adults and can lead to joint destruction and disability [1]. Genetic and environmental factors are associated with RA development [1]. Elderly patients with RA include those who present with young-onset RA (YORA) earlier in life and those with late-onset RA (LORA), where the disease begins after the age of 60-65, depending on the definition [2, 3]. The prevalence of LORA has been increasing worldwide, especially in Japan, due to accelerated global aging, especially among developed countries [4–6].

Previous studies have suggested that genetic and environmental factors may differ between patients with LORA and those with YORA. LORA is characterized by higher prevalence in males, acute onset, large joint involvement, and lower prevalence of positivity of rheumatoid factor (RF) or anti-cyclic citrullinated peptide antibody (ACPA), in comparison with YORA [6, 7]. The *HLA-DRB1* allele, such as shared epitope alleles, is the strongest genetic risk factor for RA; several studies have shown different associations of *HLA-DRB1* alleles between LORA and YORA [8]. Regarding environmental factors, current LORA patients may have been exposed to smoking for a longer period than YORA patients, given that the prevalence of smoking, one of the major environmental factors for RA, has been decreasing worldwide over the last 20 years (data from the World Health Organization; https://www.who.int).

Mosaic chromosomal alterations (mCAs) are clonal cellular expansions associated with somatic mutations, particularly large-scale chromosomal rearrangements [9]. mCAs can occur in autosomes and sex chromosomes, and their impact on health outcomes appears to differ between autosomal and sex chromosomal mCAs. Previous studies have found that autosomal mCAs are associated with haematological malignancies and aging disorders, such as infections and cardiovascular diseases [10–12]. Sex chromosomal mCAs include mosaic loss of Y (mLOY) in males and mosaic loss of X (mLOX) in females. mLOY is a highly polygenic trait associated with multiple age-related diseases such as malignancy and cardiovascular diseases [13–15], and mLOX is associated with genes related to autoimmunity [16]. While mCAs have been discussed primarily in the context of haematological malignancies and age-related diseases [17], recent studies have reported that mCAs can also be observed in healthy individuals, especially in males and the elderly [10, 13, 16, 18, 19]. Given that mCAs accumulate with age, especially in males, and reflect the clonal expansion of dysregulated immune cells, similar to non-mCA somatic mutations [10, 20], we hypothesised that mCAs have a substantial impact on the development of LORA compared to YORA. Knowledge of the association between mCAs, including mLOX and mLOY, and RA is lacking.

Here, we analysed the associations of mCAs (autosome, mLOX, and mLOY) with RA or its subsets (LORA and YORA) in 4,466 patients with RA derived from two independent datasets. Our findings suggest that mCAs substantially affect RA development, emphasising the possible different effects of mLOY between LORA and YORA.

## METHODS

### Study Participants and study design

We recruited 4,845 RA cases and 177,298 non-RA controls from the BioBank Japan cohort (BBJ cohort [21]) and the Institute of Rheumatology, Rheumatoid Arthritis (IORRA) cohort [22], and detected mCAs in peripheral blood samples (Figure 1A). The present study comprised of two independent datasets: Set 1 (discovery dataset) and Set 2 (validation dataset). Set 1 consisted of 2,107 BBJ RA cases and 86,998 non-RA controls. Set 2 consisted of 2,359 IORRA cases and 86,998 non-RA controls (Figure 1B). The BBJ cohort included 2,452 RA patients and 177,298 non-RA disease controls with targeted diseases [21], except for RA. For subjects from the BBJ, the diagnosis of RA was made by the attending physicians as described elsewhere [21], and only subjects with definitive RA were enrolled. In the IORRA cohort, RA was diagnosed by physicians according to the 1987 ACR criteria [23] and/or the 2010 ACR/EULAR classification criteria for RA [24]. We defined LORA and YORA as RA with onset at ≧ and < 60 years of age, respectively [3]. All patients provided informed consent, and this study was approved by the local ethics committee of each institute.

**Figure 1.**
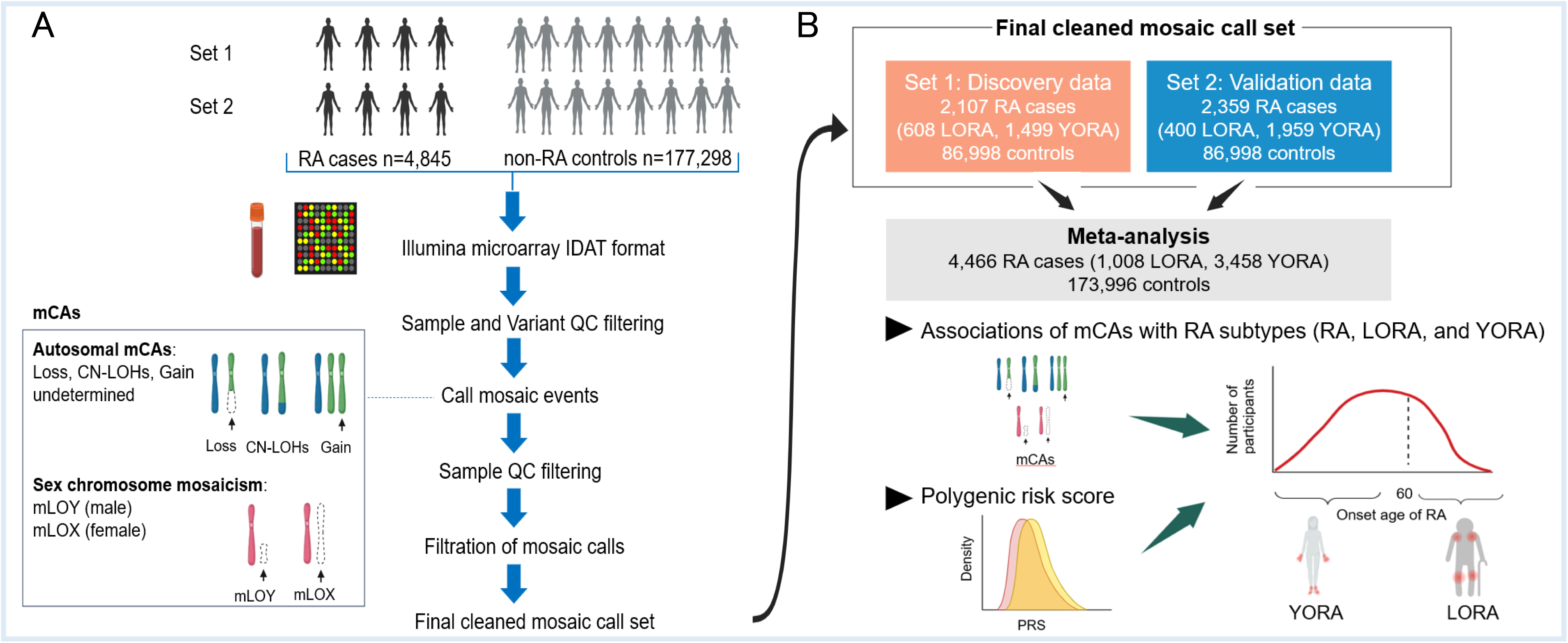
Schematic view of mosaic calling and study subjects in the current study. A. Method of mosaic calling by the MoChA. B. Study subjects in each dataset (Set 1: discovery dataset and Set 2: validation dataset). mCAs, mosaic chromosomal alterations; CN-LOHs, copy-neutral loss of heterozygosity; mLOY, mosaic loss of chromosome Y; mLOX, mosaic loss of chromosome X; LORA, late-onset RA; YORA, young-onset RA; PRS, polygenic risk score.

### Genotyping

DNA was extracted from the blood samples of all individuals. Details of genotyping of the BBJ samples have been described elsewhere [10]. Briefly, BBJ samples were genotyped using Illumina Human OmniExpress Exome v1.0 (OEEv1.0), Human OmniExpress Exome v1.2 (OEEv1.2), or a combination of Illumina HumanOmniExpress v1.0 and HumanExome v1.0, 1.1 (OE+HE). IORRA samples were genotyped using Human OmniExpress Exome v1.2.

### Variant- and Sample-level quality controls

The details of quality control (QC) steps for the variants and samples are shown in Supplementary Methods. According to the Mosaic Chromosomal Alterations (MoChA) [18, 19] protocols, we excluded variants that met any of the following criteria: genotyping call rate <97%, variants falling in segmental duplications with low divergence (<2%), variants with excess heterozygosity (p<1e-6, Hardy-Weinberg equilibrium test), and variants with allele frequency difference > 3.0% from those in the imputation panel. For the sample QCs, we removed genetically identical samples and outliers from East Asian clusters using principal component analysis (PCA). We also excluded subjects with sample call rate <0.97 and samples with phased BAF autocorrelation > 0.03.

### Detection of mosaic events

A schematic view of the detection of mosaic events is illustrated in Figure 1A, and the detailed methods are described in Supplementary Methods. Briefly, mosaic events were identified using the MoChA [18, 19]. The raw IDAT files of the subjects were used as inputs. After sample- and variant-level QC as described above, phasing was performed using SHAPEIT4 [25], and mCAs were detected. The samples were filtered after mosaic calling based on the medical history of haematological malignancies in the BBJ samples. We also removed samples that met other criteria (details are presented in Supplementary Methods). After filtering the mosaic calls, the final cleaned mosaic call set was obtained. We detected autosomal mCA events by copy number state (loss, copy-neutral loss of heterozygosity (CN-LOH), or gain) and by p versus q arm for loss and CN-LOH events. mCAs for which we could not determine the copy number state were categorised as undetermined. In addition to loss, CN-LOHs, and gain events, we included undetermined calls in the presence of autosomal mCAs. mLOY and mLOX were detected in males and females, respectively. For each mLOY that passed quality control, the cell fraction (CF) was calculated as 4*bdev/(1+2*bdev), where bdev represents the BAF deviation estimated from 0.5.

### Construction of polygenic risk score for RA

To evaluate the difference in genetic impact and effect sizes of mLOY on RA subsets (RA, LORA, and YORA) in males, we constructed a polygenic risk score (PRS). We previously conducted a multi-ancestry GWAS meta-analysis for RA [26] and constructed the best-performing PRS model, which was based on a multi-ancestry GWAS and included variants for the binding motif of T-bet in CD4^+^T cells annotated by IMPACT [27]. The best-performing model was based on 2,576 autosomal variants outside the HLA region. The imputed genotypes were used to calculate the PRS. For SNP imputation, we conducted SNP QC by excluding those with genotyping call rate < 0.97, hardy-Weinberg equilibrium p-value (HWE-P) < 1.0×10^-6^, or minor allele frequency (MAF) < 0.005. We also excluded variants with MAFs > 3.0% different from those of the imputation panel to achieve high imputation accuracy. Variants with low imputation accuracy (R^2^ < 0.3) were excluded. For an imputation panel, we used a previously constructed imputation panel from the phase 3 1000 genome project ver.5 (1KGP3ver5) (http://www.internationalgenome.org/data) [28] combined with high-depth whole genome sequencing (WGS) data from 3265 Japanese subjects from BBJ (J3K), which enables rare variant detection [29]. We extracted the SNPs shared among BBJ, IORRA, and the best-performing model (2,046 SNPs). The effect size estimates of the risk alleles from the best-performing model were then multiplied by the corresponding genotype dosages of the risk alleles and summed for each individual. We calculated the PRS for subjects who passed the sample-level QC as described above.

### Statistical analysis

We performed logistic regression analyses to evaluate the association between RA and detectable mosaicism as follows:

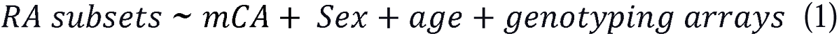

where the RA subsets are RA, LORA, and YORA, and sex, age, and types of genotyping array (OEEv1.0, OEEv1.2, and OE+HE) are the covariate terms. We separately assessed the following mosaic events: autosomal mCAs, mLOX, and mLOY. Only female and male samples were analysed for mLOX and mLOY, respectively, without sex as a covariate. We also analysed samples for which smoking information was available, adding smoking history (never or previous/current) to the covariates, and compared the results with those without smoking information. We further assessed the associations between RA subsets and the CF of mLOY (mLOY-CF) >5% in the same manner as described above to evaluate whether the associations between RA subsets and mLOY increased in a CF-dependent manner. We also assessed the association between age at disease onset and mLOY among RA subjects adjusted for age, age^2^, and array types.

Associations between genetic risk factors for RA and RA subsets in the presence of mLOY were examined in each dataset using logistic regression models adjusted for age using the following formula:

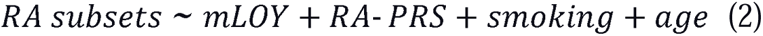

where the PRS for RA (RA-PRS) and smoking history were included as risk factors. To avoid overfitting, we removed first- or second-degree relatives showing PIHAT > 0.25 by identity-by-descent (IBD) detection. We conducted this analysis on the RA, LORA, and YORA groups. We also assessed the association between mLOY and PRS, adjusting for age, age^2^, and smoking status, to ensure no associations between them.

Furthermore, we evaluated the impact of risks on LORA by referring to YORA and applying the same logistic regression models to RA subjects. Only male participants with available smoking information were included in the three analyses to evaluate the association between PRS and mLOY.

We investigated the interactive association between mLOY and RA-PRS in male patients with LORA using a logistic regression model. First- or second-degree relatives with PIHAT > 0.25 were excluded from this analysis. We stratified the patients into six subgroups according to the PRS and mLOY status. The PRS was divided into low (<25%) and high (≧25%) based on the distribution of PRS across the RA subjects from both datasets. We set three mLOY statuses, mLOY negative, mLOY positive, and mLOY-CF >5%. Setting low genetic risk subgroups, which did not have mLOY, as a reference, we analysed the risk of LORA in each group adjusted for age in each dataset using logistic regression models and combined the results by adding the dataset (Set 1 or Set 2) as a covariate. We did not include smoking as a covariate to maximise the sample size. We assessed the interactive association between mLOY and PRS among male patients with YORA in the same manner. To further evaluate the interaction between mLOY-CF >5% and RA-PRS, we compared the goodness of fit between the following two logistic regression models using one-way ANOVA:

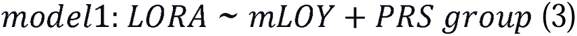

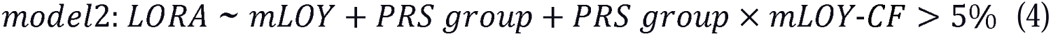

where LORA (0: non-RA, 1: LORA), PRS group (0: low, 1: high), and mLOY-CF >5% (0: mLOY-negative or mLOY-CF <5%, 1: mLOY-CF >5%) are all binary traits; hence, is an interaction term (PRS group × mLOY-CF >5 %) in Equation (4).

All statistical analyses were conducted using the R software (version 3.3.3). All analyses, except for the interactive association between mLOY and PRS, were performed separately in the two datasets (Sets 1 and 2), and the results were meta-analysed. Meta-analyses were performed using an inverse variance fixed-effects model using the metafor: Meta-Analysis Package for R. We set non-RA subjects as a reference in the analyses to evaluate the associations between mCAs or mLOY-CF >5% with RA subsets (RA, LORA, and YORA), and the associations between mLOY and risk factors for RA. The statistical significance of the associations between mCAs or mLOY-CF >5% and RA subsets (RA, LORA, and YORA) was based on the Bonferroni correction (P<0.05/3 = 0.016). The statistical significance of the interactive associations between mLOY and PRS among the five subgroups was also based on Bonferroni correction (P<0.05/5 = 0.010). In other logistic regression models, we set the significance threshold at a two-sided P-value of <0.05.

### Patient and public involvement statement

Patients were not involved in any part of this research, including the study design, formal analyses, result interpretation, writing, or editing of the manuscript for readability or accuracy.

## RESULTS

### Characteristics of study participants

The demographic features of the participants in Sets 1 (discovery dataset) and 2 (validation dataset) are listed in Table 1. Patients with RA were younger than controls in both datasets. Female sex rates were higher in patients with RA than in controls in both datasets (Set 1: 81.5% vs. 45.6%; Set 2: 85.5% vs. 45.6%). The age of patients with LORA was higher than that of the controls. In contrast, the age of patients with YORA was lower than that of controls in both datasets (Supplementary Tables S1 and S2).

**Table 1.**
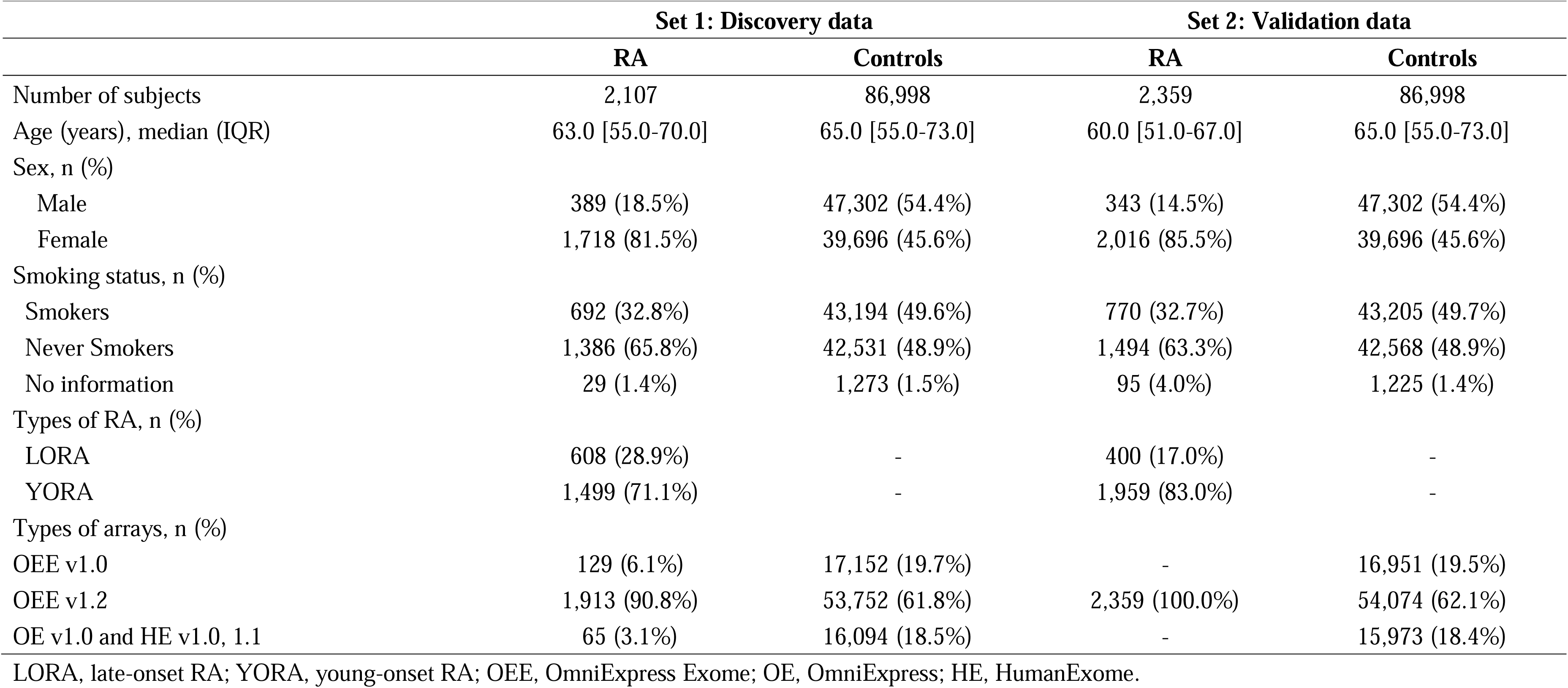
Demographic features of subjects in each dataset.

|  | Set 1: Discovery data |  | Set 2: Validation data |  |
| --- | --- | --- | --- | --- |
|  | RA | Controls | RA | Controls |
| Number of subjects | 2,107 | 86,998 | 2,359 | 86,998 |
| Age (years), median (IQR) | 63.0 [55.0-70.0] | 65.0 [55.0-73.0] | 60.0 [51.0-67.0] | 65.0 [55.0-73.0] |
| Sex, n (%) |  |  |  |  |
| Male | 389 (18.5%) | 47,302 (54.4%) | 343 (14.5%) | 47,302 (54.4%) |
| Female | 1,718 (81.5%) | 39,696 (45.6%) | 2,016 (85.5%) | 39,696 (45.6%) |
| Smoking status, n (%) |  |  |  |  |
| Smokers | 692 (32.8%) | 43,194 (49.6%) | 770 (32.7%) | 43,205 (49.7%) |
| Never Smokers | 1,386 (65.8%) | 42,531 (48.9%) | 1,494 (63.3%) | 42,568 (48.9%) |
| No information | 29 (1.4%) | 1,273 (1.5%) | 95 (4.0%) | 1,225 (1.4%) |
| Types of RA, n (%) |  |  |  |  |
| LORA | 608 (28.9%) | - | 400 (17.0%) | - |
| YORA | 1,499 (71.1%) | - | 1,959 (83.0%) | - |
| Types of arrays, n (%) |  |  |  |  |
| OEE v1.0 | 129 (6.1%) | 17,152 (19.7%) | - | 16,951 (19.5%) |
| OEE v1.2 | 1,913 (90.8%) | 53,752 (61.8%) | 2,359 (100.0%) | 54,074 (62.1%) |
| OE v1.0 and HE v1.0, 1.1 | 65 (3.1%) | 16,094 (18.5%) | - | 15,973 (18.4%) |

LORA, late-onset RA; YORA, young-onset RA; OEE, OmniExpress Exome; OE, OmniExpress; HE, HumanExome.

Consistent with previous findings [6, 7], the prevalence of LORA was higher in males, and the positivity of autoantibodies (RF or ACPA) was lower in LORA than in YORA in both datasets (Supplementary Table S3).

### Significant negative associations of autosomal mCAs and mLOX with RA

Consistent with previous studies, the prevalence of autosomal mCA events increased with age in patients with RA and controls in both datasets (Figure 2). We found that autosomal mCAs showed consistent negative associations with RA and its subsets in both datasets, even after adjusting for covariates, including age, and significant associations were confirmed by meta-analyses (Table 2 and Figures 2 and 3).

**Figure 2.**
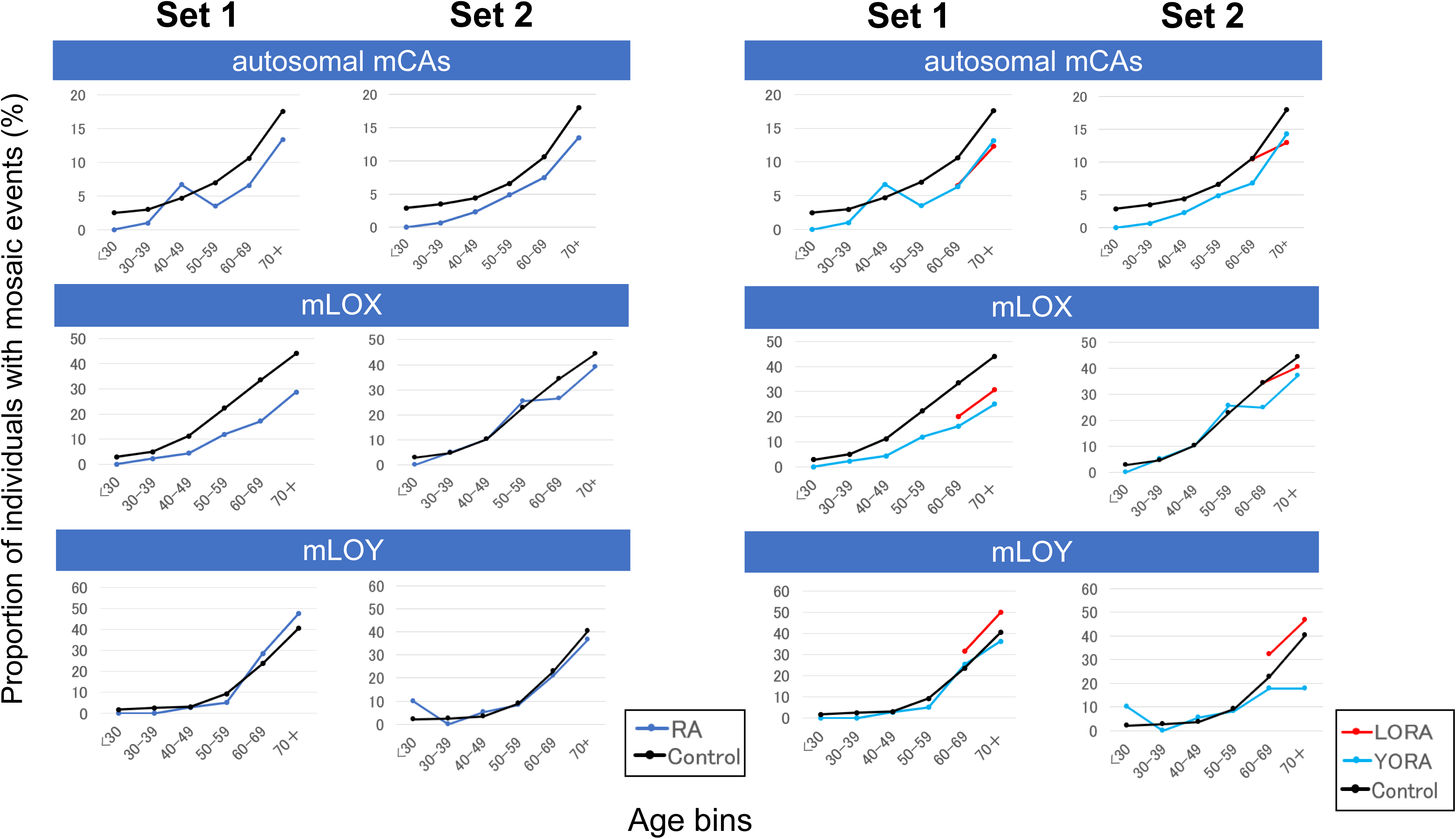
Frequency of detectable mosaic events stratified by age. The frequencies (means) are indicated for Set 1 (2,107 RA and 86,998 controls) and Set 2 (2,359 RA and 86,998 controls). mCAs, mosaic chromosomal alterations; mLOX, mosaic loss of chromosome X; mLOY, mosaic loss of chromosome Y; LORA, late-onset RA; YORA, young-onset RA.

**Figure 3.**
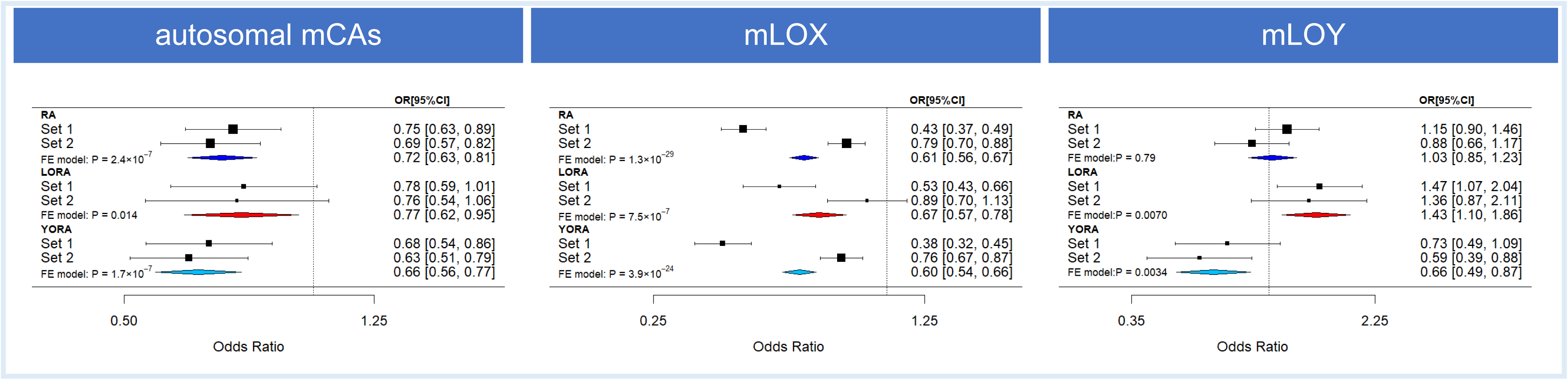
Associations between autosomal mCAs, mLOX, or mLOY and RA subsets. Associations between detectable mosaicism (autosomal mCAs, mLOY, and mLOX) and RA subsets (RA, LORA, and YORA) were evaluated by comparison with non-RA controls. Forest plots of the meta-analysis for each association analysis using two datasets (Sets 1 and 2) are shown. ORs are indicated by squares and 95% CIs are indicated by two-sided lines. The corresponding data are listed in Table 2. mCAs, mosaic chromosomal alterations; mLOX, mosaic loss of chromosome X; mLOY, mosaic loss of chromosome Y; LORA, late-onset RA; YORA, young-onset RA.

**Table 2.**
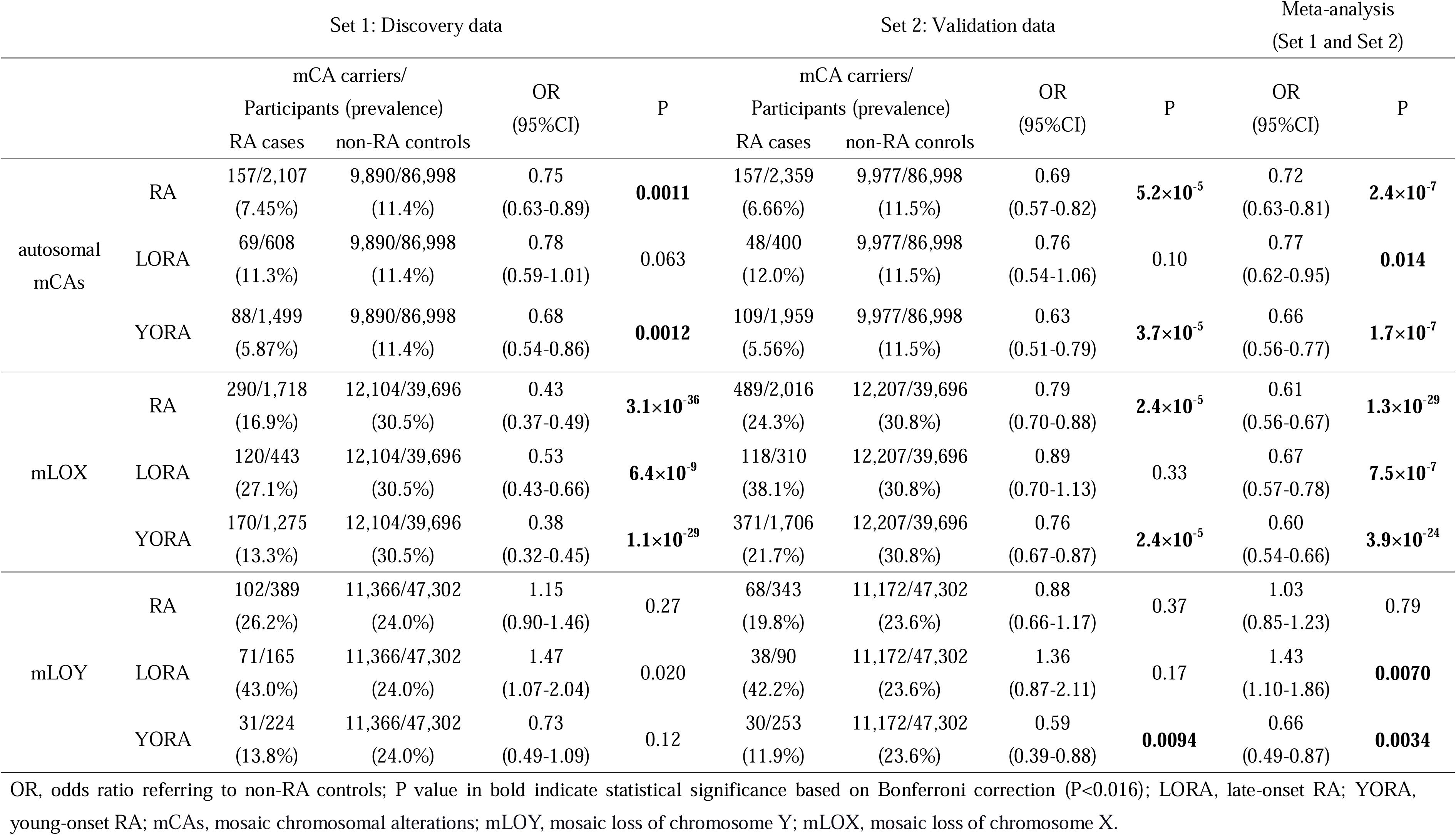
The associations between different types of mCAs and RA subsets (RA, LORA, and YORA) in each dataset corresponding to. **Figure 3**

As observed for autosomal mCAs, mLOX increased with age (Figure 2). A logistic regression model adjusted for covariates, including age, revealed consistent negative associations between mLOX and RA and its subsets, and significant associations were confirmed by the meta-analysis (Table 2 and Figures 2 and 3). As cigarette smoking is associated with mCAs [14], we included smoking history as a covariate in the statistical model, in which we confirmed significant negative associations between autosomal mCAs or mLOX and RA or its subsets, even at the expense of decreased sample sizes (Supplementary Figure S1, Table S4).

### Significant positive association between mLOY and LORA

When we analysed the association of mLOY with RA and its subsets in male samples, mLOY was positively associated with LORA in each dataset and meta-analysis. However, no consistent association was observed between mLOY and RA. Notably, we observed a protective association between mLOY and YORA, which is in stark contrast to that observed for LORA (Table 2 and Figures 2 and 3). The directions of the effect sizes of mLOY on both LORA and YORA were concordant for both datasets (Figure 3; Table 2). We confirmed robust associations in a statistical model that included smoking history as a covariate (Supplementary Figure S1, Table S4). Since the age definition of LORA varied among different populations [6], we also evaluated the association between age at RA onset and mLOY, which showed a significant positive association in the meta-analysis adjusted for covariates, including age (Supplementary Figure S2, Table S5), further supporting a positive association between mLOY and LORA, regardless of the definition of age for LORA.

### Significant and CF-dependent association between mLOY and LORA

Since the cell fraction (CF) of mLOY varies among individuals, we hypothesised that the higher the CF of mLOY, the larger the effect size of mLOY on LORA. To test this hypothesis, we extracted subjects with RA or its subsets whose CF of mLOY was > 5% (mLOY-CF >5%) (Supplementary Figure S3). We examined their association with mLOY using a logistic regression model adjusted for covariates, including age. We found that the effect sizes of mLOY-CF >5% were higher than those of mLOY without the CF threshold on LORA in both the datasets and the meta-analysis (Figure 4 and Supplementary Table S6), indicating that the effect size of mLOY on LORA increases in a CF-dependent manner. In contrast, we did not observe consistent CF-dependent changes in the effect sizes of YORA, whereas the same non-significant trends were observed in RA (Figure 4). The observed associations were not affected by the adjustment for smoking status (Supplementary Figure S4, Table S7).

**Figure 4.**
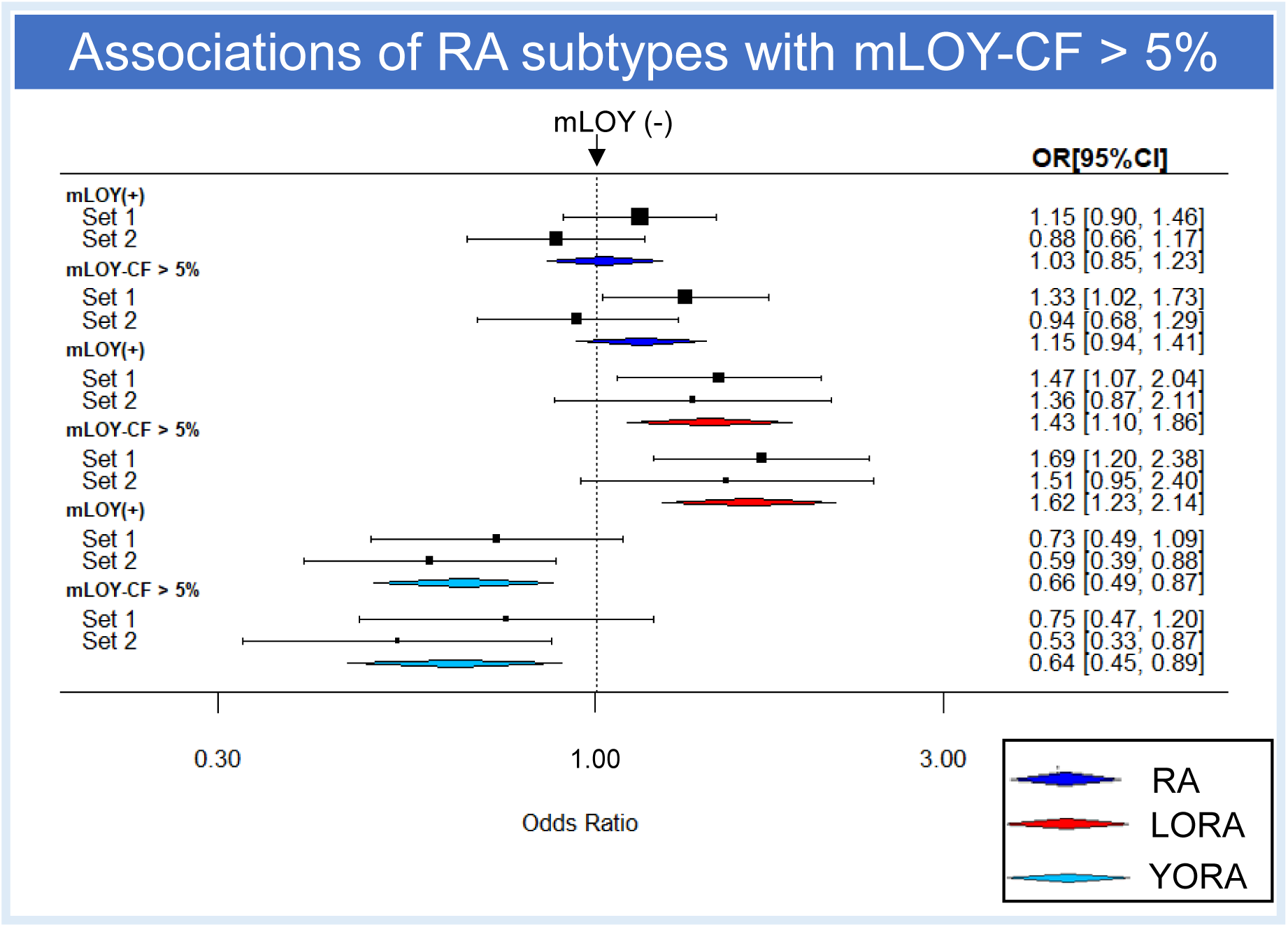
Associations of CF of mLOY >5% with RA and its subsets. Associations between mLOY-CF >5% and the RA subsets (RA, LORA, and YORA) were evaluated referring to non-RA controls. The associations of mLOY without the CF threshold with the RA subsets (RA, LORA, and YORA) are also shown. Forest plots of the meta-analysis for each association analysis using two datasets (Sets 1 and 2) are shown. ORs are indicated by squares and 95%CIs are indicated by two-sided lines. The corresponding data are listed in Supplementary Table S6. mLOY, mosaic loss of chromosome Y; LORA, late-onset RA; YORA, young-onset RA.

### Distinct associations of RA-PRS and mLOY between LORA and YORA

Polygenic score (PRS) measures the collection of associations between heritable genetic components, causal SNPs, or their proxies, while mosaic events occur after birth and accumulate with age. Accordingly, we evaluated the associations of mLOY and PRS for RA with RA subsets (RA, LORA, and YORA) using a model adjusted for smoking status in each dataset (Sets 1 and 2, Supplementary Table S8) and meta-analysis. Notably, mLOY was not associated with the PRS in patients with RA (Supplementary Table S9). The effect size of PRS was higher in the YORA group (beta = 0.61 [95%CI 0.55-0.66], P = 3.0×10^-111^) than in the LORA group (beta = 0.44 [95%CI 0.37-0.51], P = 1.7×10^-32^) in the individual datasets and meta-analysis (Figure 5A, Supplementary Table S10). We also confirmed a positive association between mLOY and LORA (beta = 0.32, P = 0.017) and a negative association between mLOY and YORA (beta = -0.41, P = 0.0047) during conditioning on the PRS for RA (Figure 5B, Supplementary Table S10). Among the RA subjects, the PRS was lower in LORA than in YORA (beta = -0.16, P = 0.014). mLOY demonstrated a borderline significant positive association with LORA compared to YORA (beta = 0.46, P = 0.067) (Supplementary Figure S5, Table S11). These data suggest distinct underlying somatic architectures between LORA and YORA, in addition to their different genetic backgrounds.

**Figure 5.**
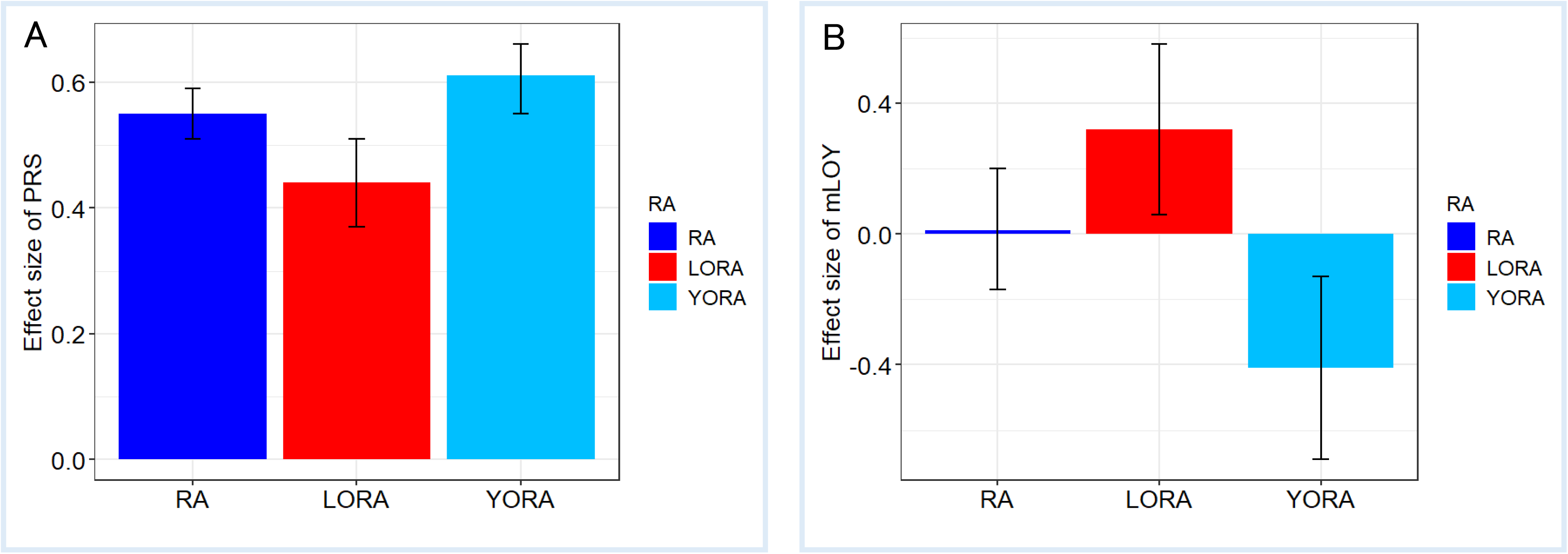
Comparison of effect sizes of PRS and mLOY among RA and its subsets. The effect sizes of the PRS (A) or mLOY (B), referring to non-RA controls, were compared among the RA subsets (RA, LORA, and YORA). The effect sizes of the PRS (A) and mLOY (B) of the meta-analysis for each association analysis using the two datasets (Sets 1 and 2) are shown. Effect sizes are indicated in vertical columns and 95%CI of effect sizes are indicated by two-sided lines. The corresponding data are presented in Supplementary Table S10. PRS, polygenic risk score; mLOY, mosaic loss of chromosome Y; LORA, late-onset RA; YORA, young-onset RA.

### Dose-dependent association of mLOY and a potential interactive association of mLOY and RA-PRS on LORA

Finally, we evaluated the dose-dependent association of mLOY and the potential interactive association between RA-PRS and mLOY in LORA and YORA (Supplementary Table S8). Patients were stratified into six subgroups based on PRS (low or high) and mLOY status (positive, negative, or mLOY-CF >5%); a subgroup of patients with low PRS and mLOY(-) was used as a reference. Among the low PRS subgroups, both the mLOY(+) and mLOY-CF >5% subgroups showed a non-significant slight increase in the risk of LORA (Figure 6A, upper panel). Interestingly, among the high-PRS subgroups, the risk of LORA sharply and significantly increased in a CF-dependent manner (Figure 6A, lower panel). In contrast, as for YORA, we observed a significant increase in the risk among high PRS subgroups, but in a CF-independent manner or rather in a trend of a CF-dependent protective manner (Figure 6B, lower panel), highlighting a different association of mLOY-CF on LORA and YORA. These associations were consistent in each dataset (Supplementary Figures S6 and S7). To further investigate the potential interaction between RA-PRS and mLOY-CF >5% in LORA, we introduced an interaction term (PRS group × mLOY-CF >5%) into the model. We compared the goodness of fit between the models with and without the interaction term using a one-way ANOVA. We found that the model with the interaction term fit significantly better than the model without the interaction term (P = 0.0036) (Supplementary Table S12), supporting a possible interaction between mLOY-CF >5% and the PRS in the LORA.

**Figure 6.**
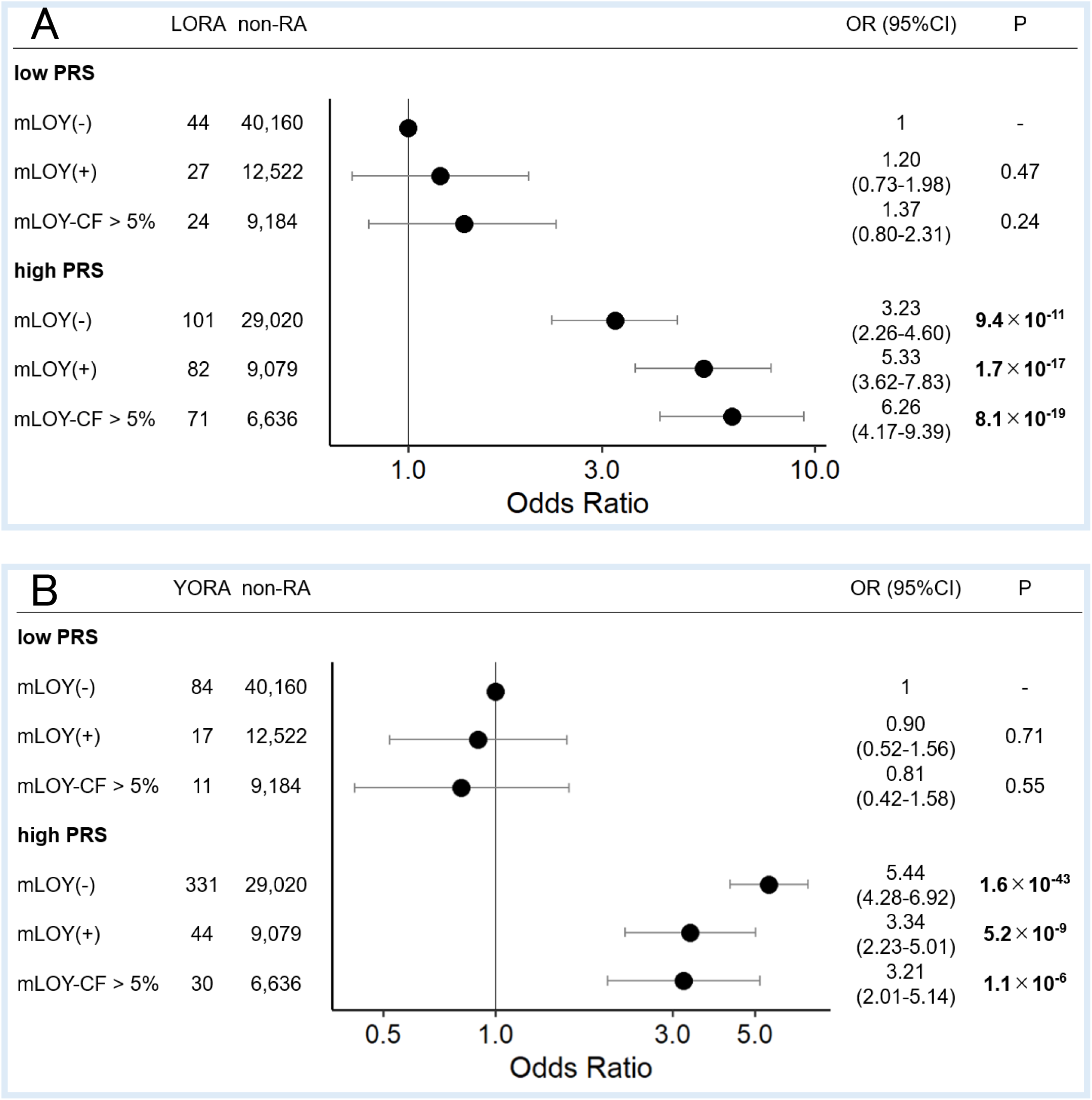
Combinatory associations of PRS and mLOY on LORA and YORA. The associations of the PRS subgroups with LORA or YORA stratified by mLOY status were evaluated referring to low PRS subgroup without mLOY. ORs are indicated by dots, and 95%CIs are indicated by two-sided lines. PRS, polygenic risk score; mLOY, mosaic loss of chromosome Y; LORA, late-onset RA; YORA, young-onset RA.

## DISCUSSION

The association between somatic mutations in peripheral blood mononuclear cells (PBMCs) and RA has been previously investigated. Savola et al. explored a possible association between clonal haematopoiesis of indeterminate potential (CHIP) and the expansion of cells with somatic point mutations. However, a significant association between CHIP and RA was not found [30]. In contrast, our previous study observed a trend toward a protective association between RA and autosomal mCAs [10]. In the present study, we investigated the association between mCAs and RA or its subsets (LORA and YORA) and found that mLOY was positively associated with LORA, while mLOY was negatively associated with YORA. We also found consistent negative associations between autosomal mCAs (as reported in our previous study [10]) and mLOX in RA, LORA, and YORA. Although PRS was weakly associated with LORA compared to YORA, the association between PRS and LORA was augmented by mLOY in a dose-dependent manner. In contrast, the positive associations between PRS and YORA were independent of mLOY or showed a trend of protective associations in the presence of mLOY.

It is well known that genetic and environmental factors, as well as their interaction, play significant roles in the development of RA [1]. Previous studies have identified both genetic and environmental risks of RA susceptibility, seropositivity of autoantibodies (RF or ACPA), and joint destruction [1]. However, the target samples in most studies were YORA subjects; hence, neither the genetic and environmental risks specific to LORA have been thoroughly investigated. The smaller effect sizes of PRS in LORA than in YORA observed in the present study suggest that the heritable components of disease risk, or at least their contributions, differ between LORA and YORA. Regarding environmental (somatic) components in LORA, acquired age-related alterations of the immune system, including immunosenescence and inflammaging, have recently been implicated in the development of LORA [31]. We hypothesised that acquired immune alterations could be caused by mCAs and their subsequent gene-environment interactions. The observed significant positive association between mLOY and LORA, along with the accumulation of mLOY with age and the possible interaction between PRS and mLOY, supports the idea that mLOY may play a pivotal role in the development of LORA. Indeed, recent studies have found that in addition to the heritable germline variants, the acquired somatic mutations lead to the development of autoimmune and autoinflammatory diseases later in life [32, 33]. Overall, these results suggest that the underlying genetic and environmental (somatic) architecture of the LORA is distinct from that of the YORA.

As for the biological mechanisms, mLOY is known to be associated with age-related disorders, such as cardiovascular disease or Alzheimer’s disease [15, 34], by directly altering gene functions in particular immune cell subsets [35] or causing genetic instability, leading to clonal expansion of dysregulated immune cells [13, 36]. Hence, LORA may also develop in dysregulated immune systems as a consequence of the accumulation of mLOY in certain immune cell types. In contrast, we observed consistent protective associations between autosomal mCAs, mLOX, and RA. Future studies, such as single-cell RNA sequencing of patient-derived immune cells, will clarify the downstream effects of mLOY in a cell type-specific manner.

Several epidemiological studies have shown an increase in the prevalence of LORA in recent years [5, 6], reflecting the acceleration of aging societies, especially in developed countries. Males are more susceptible to LORA, whereas females are more likely to develop autoimmune diseases in general, including YORA [37], probably due to the effects of estrogen or progesterone on immune cells [38]. The significant positive association between mLOY and LORA partly but reasonably explains the male predominance of LORA susceptibility. Since not all women are protected from LORA, the male predominance needs to be further addressed in future studies.

Our study had several limitations. First, the sample size was limited, especially for male patients with LORA. At this point, most of the analyses were conducted separately in independent datasets, which were then meta-analysed to evaluate robustness. Although we admit that not all the tests fulfilled the stringent thresholds, such as Bonferroni-corrected p-values, we still observed similar trends in individual datasets, even with relatively small sample sizes. Second, as we did not test the causality of mLOY on LORA or vice versa owing to the sample size limitation, causality needs to be tested by enrolling more subjects to enable us to construct a reliable PRS specifically for LORA. Finally, the association between PRS and RA might be inflated owing to partial sample overlap between the subjects in the present study and those in the previous GWAS [26]. While we warrant testing the replicability of the datasets completely independent of the previous GWAS, the aim of this analysis was not to evaluate the PRS performance; thus, the impact of sample overlap on our analyses would be minimal, if any.

To the best of our knowledge, this is the first study to demonstrate distinct associations of mCAs between LORA and YORA, which may explain the different pathophysiological mechanisms and clinical manifestations between the RA subsets.

## Competing Interests

S.U., Y.I., K.I., S.H., K.H., E.T., and C.T. have no conflicts of interest.

Y.K. has research grants from Takeda Pharmaceuticals.

T.G. received speaking fees from Asahi Kasei Pharma, Astellas, Boehringer-Ingelheim, Bristol-Myers Squibb, Chugai, Eisai, Eli Lily, Gilead Sciences, Ono Pharmaceuticals, Pfizer, and Tanabe Mitsubishi.

G.G. declared competing interests, and patent application PCT/WO2019/079493 has been filed for the mosaic chromosomal alteration detection method used in this study.

M.K. has received consulting fees, speaking fees, and/or research grants from Argenx, Asahi Kasei Parma, AstraZeneca, Boehringer Ingelheim, Chugai, GSK, Janssen, Kissei, MBL, Mochida, MSD, Novartis, Ono Pharmaceuticals, and Tanabe Mitsubishi.

## Acknowledgement

This work was supported by the Japan Agency for Medical Research and Development (AMED) grants 21ek0109555, 21tm0424220, 21ck0106642, 23ek0410114, and 23tm0424225; the Japan Society for the Promotion of Science (JSPS) KAKENHI grant JP20H00462; and Takeda Hosho Grants for Research in Medicine. The cartoons shown in Fig. 1 were created using Bio-Render. com. We thank the staff of BBJ for collecting and managing the samples and clinical information.

## Author contributions

M.K. and C.T. conceived the project. S.U. and C.T. analysed the data. K.H. shared the statistical codes. K. I., S. H., and E. T. generated the IORRA data. G.G. developed MoChA pipelines for mosaic calling. S.U., Y.I., and C.T. wrote the manuscript. All authors have critically reviewed and approved the final version of the manuscript.

## Data availability

Statistical codes are available from Chikashi Terao (ORCID 0000-0002-6452-4095) at any time, only on reasonable request. The Mosaic Chromosomal Alterations (MoChA) pipelines used for mosaic calling (mocha.wdl) are available at (https://github.com/freeseek/mochawdl).

## Notes

### Author Declarations

the ethics committees of RIKEN Center for Integrative Medical Sciences gave ethical approval for this work.

## REFERENCES

1. Rheumatoid arthritis. Nat Rev Dis Primers 2018;4:18002.

2. Kobak S, Bes C. An autumn tale: geriatric rheumatoid arthritis. Ther Adv Musculoskelet Dis 2018;10:3–11.

3. Deal CL, Meenan RF, Goldenberg DL, et al. The clinical features of elderly-onset rheumatoid arthritis. A comparison with younger-onset disease of similar duration. Arthritis Rheum 1985;28:987–94.

4. Collaborators GBDRA. Global, regional, and national burden of rheumatoid arthritis, 1990-2020, and projections to 2050: a systematic analysis of the Global Burden of Disease Study 2021. Lancet Rheumatol 2023;5:e594–610.

5. Kato E, Sawada T, Tahara K, et al. The age at onset of rheumatoid arthritis is increasing in Japan: a nationwide database study. Int J Rheum Dis 2017;20:839–45.

6. Uchiyama S, Takanashi S, Takeno M, et al. Should we reconsider the definition of elderly-onset rheumatoid arthritis in an ageing society? Mod Rheumatol 2022;32:323–9.

7. Serhal L, Lwin MN, Holroyd C, et al. Rheumatoid arthritis in the elderly: Characteristics and treatment considerations. Autoimmun Rev. 2020;19:102528.

8. Gonzalez-Gay MA, Hajeer AH, Dababneh A, et al. Seronegative rheumatoid arthritis in elderly and polymyalgia rheumatica have similar patterns of HLA association. J Rheumatol 2001;28:122–5.

9. Liu X, Kamatani Y, Terao C. Genetics of autosomal mosaic chromosomal alteration (mCA). J Hum Genet 2021;66:879–85.

10. Terao C, Suzuki A, Momozawa Y, et al. Chromosomal alterations among age-related haematopoietic clones in Japan. Nature 2020;584:130–5.

11. Zekavat SM, Lin SH, Bick AG, et al. Hematopoietic mosaic chromosomal alterations increase the risk for diverse types of infection. Nat Med 2021;27:1012–24.

12. Evans MA, Walsh K. Clonal hematopoiesis, somatic mosaicism, and age-associated disease. Physiol Rev 2023;103:649–716.

13. Thompson DJ, Genovese G, Halvardson J, et al. Genetic predisposition to mosaic Y chromosome loss in blood. Nature 2019;575:652–7.

14. Terao C, Momozawa Y, Ishigaki K, et al. GWAS of mosaic loss of chromosome Y highlights genetic effects on blood cell differentiation. Nat Commun 2019;10:4719.

15. Sano S, Horitani K, Ogawa H, et al. Hematopoietic loss of Y chromosome leads to cardiac fibrosis and heart failure mortality. Science 2022;377:292–7.

16. Liu A, Genovese G, Zhao Y, et al. Population analyses of mosaic X chromosome loss identify genetic drivers and widespread signatures of cellular selection. medRxiv. 2023 Jan 31.

17. Laurie CC, Laurie CA, Rice K, et al. Detectable clonal mosaicism from birth to old age and its relationship to cancer. Nat Genet 2012;44:642–50.

18. Loh PR, Genovese G, McCarroll SA. Monogenic and polygenic inheritance become instruments for clonal selection. Nature 2020; 584:136–41.

19. Loh PR, Genovese G, Handsaker RE, et al. Insights into clonal haematopoiesis from 8,342 mosaic chromosomal alterations. Nature 2018;559:350–5.

20. Alriyami M, Polychronakos C. Somatic Mutations and Autoimmunity. Cells 2021;10:2056. 10.3390/cells10082056

21. Nagai A, Hirata M, Kamatani Y, et al. Overview of the BioBank Japan Project: Study design and profile. J Epidemiol 2017; 27:S2–8.

22. Yamanaka H, Tanaka E, Nakajima A, et al. A large observational cohort study of rheumatoid arthritis, IORRA: Providing context for today’s treatment options. Mod Rheumatol. 2020;30:1–6.

23. Arnett FC, Edworthy SM, Bloch DA, et al. The American Rheumatism Association 1987 revised criteria for the classification of rheumatoid arthritis. Arthritis Rheum 1988;31:315–24.

24. Aletaha D, Neogi T, Silman AJ, et al. 2010 rheumatoid arthritis classification criteria: an American College of Rheumatology/European League Against Rheumatism collaborative initiative. Ann Rheum Dis 2010;69:1580–8.

25. Delaneau O, Zagury JF, Robinson MR, et al. Accurate, scalable and integrative haplotype estimation. Nat Commun 2019;10:5436.

26. Ishigaki K, Sakaue S, Terao C, et al. Multi-ancestry genome-wide association analyses identify novel genetic mechanisms in rheumatoid arthritis. Nat Genet 2022;54:1640–51.

27. Amariuta T, Ishigaki K, Sugishita H, et al. Improving the trans-ancestry portability of polygenic risk scores by prioritizing variants in predicted cell-type-specific regulatory elements. Nat Genet 2020;52:1346–54.

28. Genomes Project C, Auton A, Brooks LD, et al. A global reference for human genetic variation. Nature 2015;526:68–74.

29. Ito S, Liu X, Ishikawa Y, et al. Androgen receptor binding sites enabling genetic prediction of mortality due to prostate cancer in cancer-free subjects. Nat Commun 2023;14:4863.

30. Savola P, Lundgren S, Keranen MAI, et al. Clonal hematopoiesis in patients with rheumatoid arthritis. Blood Cancer J 2018;8:69.

31. Bauer ME. Accelerated immunosenescence in rheumatoid arthritis: impact on clinical progression. Immun Ageing 2020; 17:6.

32. Sikora KA, Wells KV, Bolek EC, et al. Somatic mutations in rheumatological diseases: VEXAS syndrome and beyond. Rheumatology (Oxford) 2022;61:3149–60.

33. Beck DB, Ferrada MA, Sikora KA, et al. Somatic Mutations in UBA1 and Severe Adult-Onset Autoinflammatory Disease. N Engl J Med 2020;383:2628–38.

34. Dumanski JP, Lambert JC, Rasi C, et al. Mosaic Loss of Chromosome Y in Blood Is Associated with Alzheimer Disease. Am J Hum Genet 2016; 98:1208–19.

35. Dumanski JP, Halvardson J, Davies H, et al. Immune cells lacking Y chromosome show dysregulation of autosomal gene expression. Cell Mol Life Sci 2021; 78:4019–33.

36. Guo X, Dai X, Zhou T, et al. Mosaic loss of human Y chromosome: what, how and why. Hum Genet 2020; 139:421–46.

37. Klein SL, Flanagan KL. Sex differences in immune responses. Nat Rev Immunol 2016; 16:626–38.

38. Moulton VR. Sex Hormones in Acquired Immunity and Autoimmune Disease. Front Immunol 2018; 9:2279.

