## Supplementary for "Mosaic loss of chromosome Y characterizes late-onset rheumatoid arthritis and contrasting associations of polygenic risk score based on age at onset"

**Supplementary Methods**

**Variant- and Sample- level quality controls**

We performed variant- and sample-level quality control (QC) steps before phasing for calling of mosaic events. According to the Mosaic Chromosomal Alterations (MoChA) WDL [1,2] protocols, we excluded variants that met any of the following criteria: genotyping call rate <97%, variants falling in segmental duplications with low divergence (<2%), variants with excess heterozygosity (p<1e-6, Hardy-Weinberg equilibrium test), variants whose allele frequency difference from those of the imputation panel > 3.0%. For a imputation panel, we used a previously constructed imputation panel from the phase 3 1000 genome project ver.5 (1KGP3ver5) (<http://www.internationalgenome.org/data>) [3] combined with high-depth WGS data from 3265 Japanese subjects from BBJ (J3K), which enables rare variant detection [4]. For the QC of samples, we removed genetically identical samples showing PIHAT > 0.9 estimated by PLINK 1.9 [5] and outliers from East Asian clusters after we mapped the BBJ and IORRA samples with 1000 Genome projects [6] using principal component analysis (PCA). We also removed subjects with sample call rate < 0.97 and samples with phased BAF autocorrelation > 0.03, which is indicative of poor quality or contamination of DNA.

**Detection of mosaic events**

The detection of mCAs among the RA cases and non-RA controls was conducted on raw IDAT files. Firstly, the raw IDAT files of the subjects were used as inputs of MoChA [1,2]. The MoChA pipeline first used Illumina’s Gencall algorithm to normalize and call genotypes from raw intensities, and then converted Illumina genotyping data to VCF files using the bcftools gtc2vcf plugin. Genotype intensities were converted to BAF (B allele frequency) and LRR (Log-R ratio). After sample- and variant-level QC, phasing was performed by SHAPEIT4 [7] using all samples together. Then, the mCAs were detected by the long-range haplotype phasing method using MoChA WDL pipelines. After the MoChA, we excluded samples who met one of the following criteria: samples less than 16 years old at genotyping, samples with genotype and phenotype sex discordance, and samples with missing data on their sex and/or age status. We also removed samples with a past medical history of haematological malignancies only in the BBJ samples because we observed a strong association between autosomal mCAs and haematological malignancy [8]. We included RA samples if the onset age of RA was over 16 years. The filtering of mosaic calls was set as follows, and mosaic chromosomal alterations that have met one of these criteria were removed: germline copy number polymorphism (CNPs), constitutional mosaic events (lod_baf_phase <10), and germline duplications (length < 500 kbp and relative coverage > 2.5). To filter out XXY and XXX samples, we restricted mLOX and mLOY with estimated ploidy less than 2.5. Finally, we detected autosomal mCA events by copy-number state (loss, copy-neutral loss of heterozygosity (CN-LOH), or gain) and by p versus q arm for loss and CN-LOH events. mCAs for which we could not determine the copy number state were categorised as undetermined. In addition to loss, CN-LOHs, and gain events, we also included undetermined calls in the presence of autosomal mCAs. mLOY and mLOX were detected in males and females, respectively. The 2022-12-14 version of the MoChA WDL pipeline was used to detect autosomal mCAs, mLOX, and mLOY.

**Supplementary References**

1.	Loh PR, Genovese G, McCarroll SA. Monogenic and polygenic inheritance become instruments for clonal selection. Nature 2020; 584:136-41.

2.	Loh PR, Genovese G, Handsaker RE, et al. Insights into clonal haematopoiesis from 8,342 mosaic chromosomal alterations. Nature 2018;559:350-5.

3.	Genomes Project C, Auton A, Brooks LD, et al. A global reference for human genetic variation. Nature 2015;526:68-74.

4.	Ito S, Liu X, Ishikawa Y, et al. Androgen receptor binding sites enabling genetic prediction of mortality due to prostate cancer in cancer-free subjects. Nat Commun 2023;14:4863.

5.	Purcell S, Neale B, Todd-Brown K, et al. PLINK: a tool set for whole-genome association and population-based linkage analyses. Am J Hum Genet. 2007; 81:559-75.

6.	Genomes Project C, Abecasis GR, Altshuler D, et al. A map of human genome variation from population-scale sequencing. Nature. 2010 28; 467:1061-73.

7.	Delaneau O, Zagury JF, Robinson MR, et al. Accurate, scalable and integrative haplotype estimation. Nat Commun 2019;10:5436.

8.	Nagai A, Hirata M, Kamatani Y, et al. Overview of the BioBank Japan Project: Study design and profile. J Epidemiol 2017; 27:S2-8.

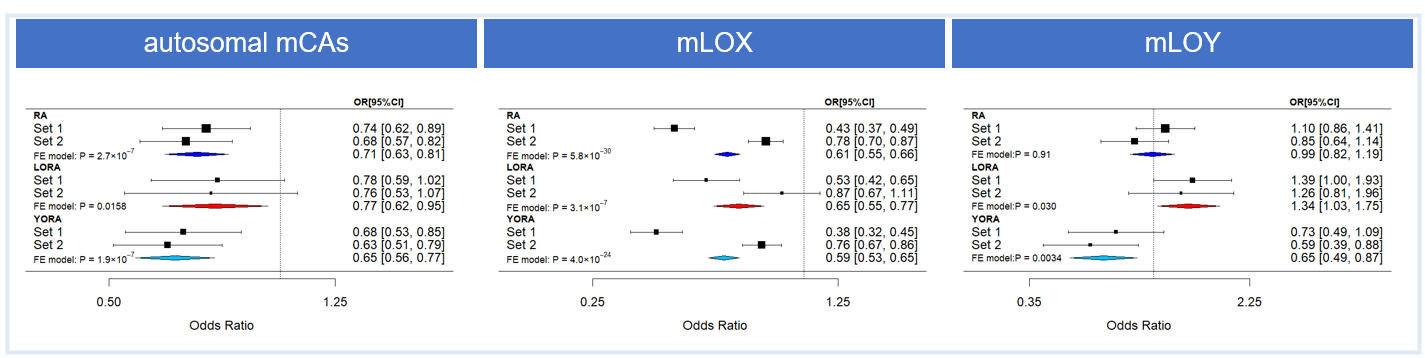

**Supplementary Figure S1. Associations between autosomal mCAs, mLOX, or mLOY and RA subsets adjusted for smoking status.**

Associations between detectable mosaicism (autosomal mCAs, mLOY, and mLOX) and RA subsets (RA, LORA, and YORA) were evaluated by comparison with non-RA controls among the subjects whose smoking information were available. Forest plots of the meta-analysis for each association analysis using two datasets (Sets 1 and 2) are shown. ORs are indicated by squares, and 95%CIs are indicated by two-sided lines. The corresponding data are listed in Supplementary Table S4. mCAs, mosaic chromosomal alterations; mLOX, mosaic loss of chromosome X; mLOY, mosaic loss of chromosome Y; LORA, late-onset RA; YORA, young-onset RA.

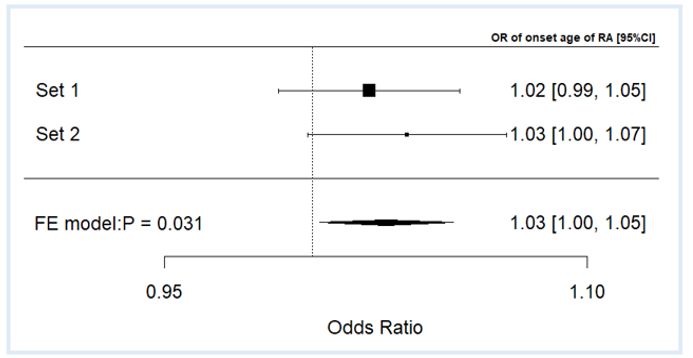

**Supplementary Figure S2. The positive association between age of RA onset and mLOY among RA subjects.**

Associations between age at RA onset and mLOY were evaluated among RA subjects. Forest plot of the meta-analysis for each association analysis using two datasets (Set 1 and Set 2) is shown. ORs are indicated by squares, and 95%CIs are indicated by two-sided lines. The corresponding data is listed in Supplementary Table S5.

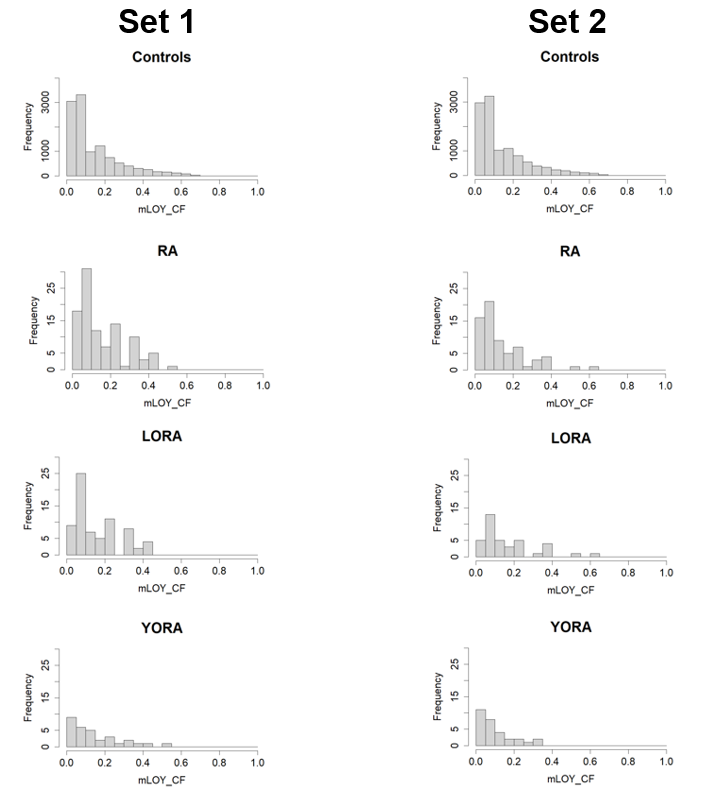

**Supplementary Figure S3. The distributions of mLOY cell fractions for mLOY positive samples in each dataset.**

The distribution of cell fraction (CF) of mLOY for 11,366 controls, 102 RA, 71 LORA and 31 YORA with mLOY events in Set 1. B. The distribution of CF of mLOY for 11,172 controls, 68 RA, 38 LORA and 30 YORA with mLOY events in Set 2. The number of samples is shown on the y-axis. mLOY, mosaic loss of chromosome Y; LORA, late-onset RA; YORA, young-onset RA.

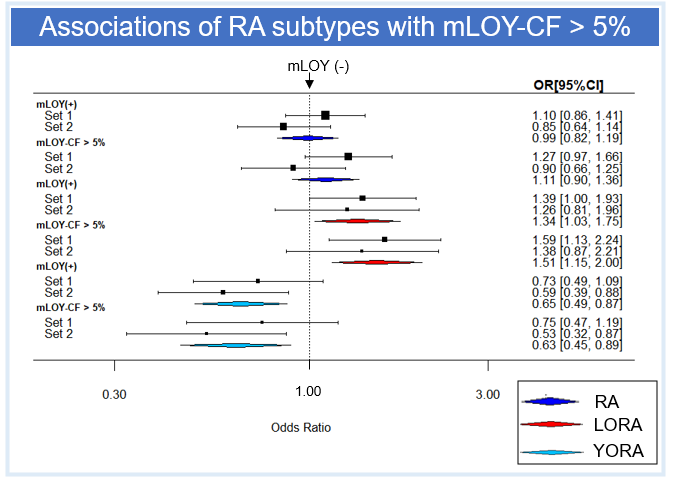

**Supplementary Figure S4. Associations of CF of mLOY >5% with RA and its subsets adjusted for smoking status.**

Associations of mLOY-CF >5% and the RA subsets (RA, LORA, and YORA) were evaluated referring non-RA controls among the subjects whose smoking information were available. The associations of mLOY without the CF threshold with the RA subsets (RA, LORA, and YORA) were also shown. Forest plots of the meta-analysis for each association analysis using two datasets (Sets 1 and 2) are shown. ORs are indicated by squares, and 95%Cis are indicated by two-sided lines. The corresponding data are listed in Supplementary Table S7. mLOY, mosaic loss of chromosome Y; LORA, late-onset RA; YORA, young-onset RA.

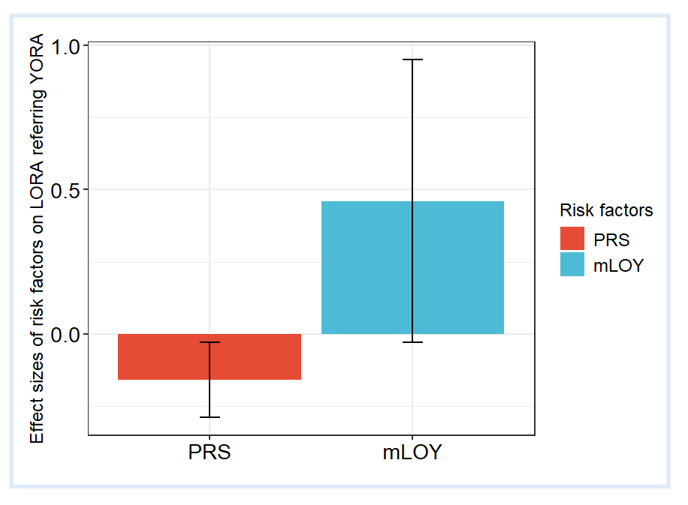

**Supplementary Figure S5. The effect sizes of PRS and mLOY on LORA referring to YORA.**

Associations of PRS or mLOY with LORA were evaluated referring to YORA. The effect sizes of the PRS or mLOY of the meta-analysis for each association analysis using two datasets (Set 1 and Set 2) are shown. Effect sizes are indicated in vertical columns and 95%CI of effect sizes are indicated by two sided lines. The corresponding data are listed in Supplementary Table S11. PRS, polygenic risk score; mLOY, mosaic loss of chromosome Y; LORA, late-onset RA; YORA, young-onset RA.

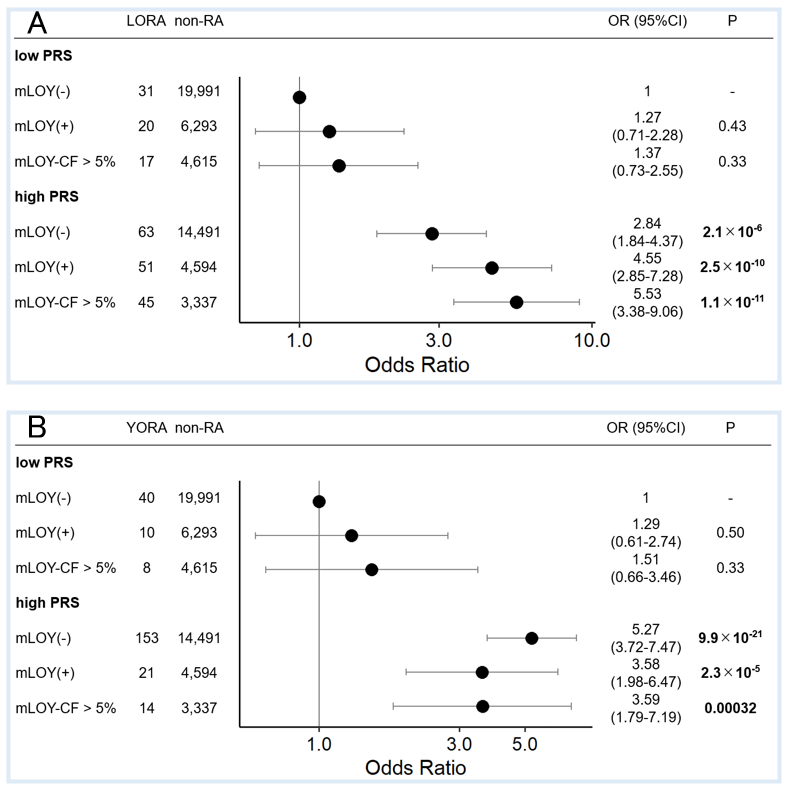

**Supplementary Figure S6. Combinatory associations of PRS and mLOY on LORA and YORA among subjects in Set 1.**

The associations of the PRS subgroups with LORA or YORA stratified by mLOY status were evaluated referring to low PRS subgroup without mLOY in Set 1. ORs are indicated by dots, and 95%CIs are indicated by two-sided lines. PRS, polygenic risk score; mLOY, mosaic loss of chromosome Y; LORA, late-onset RA; YORA, young-onset RA.

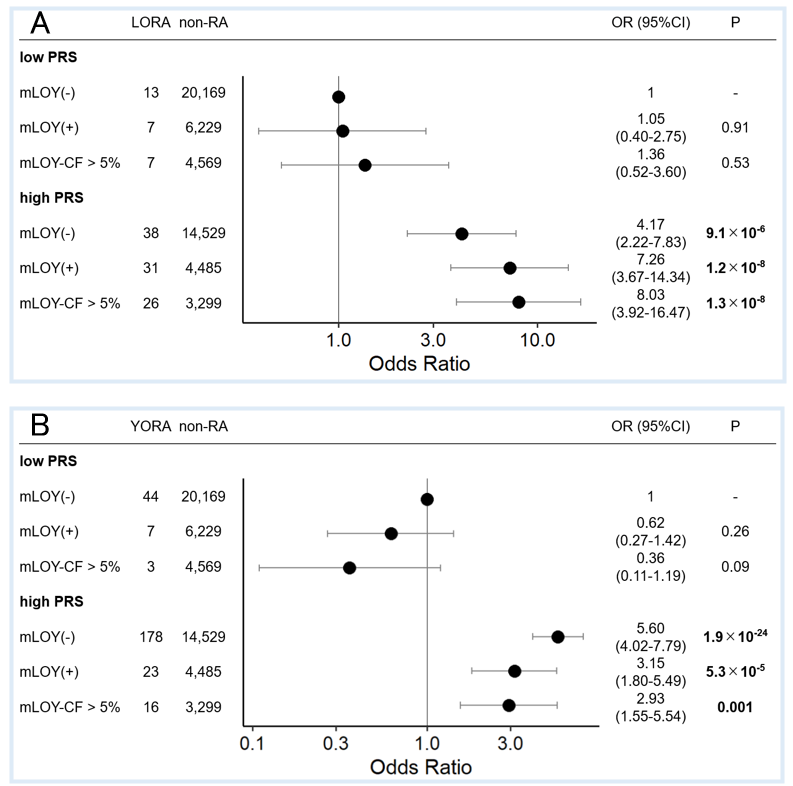

**Supplementary Figure S7. Combinatory associations of PRS and mLOY on LORA and YORA among subjects in Set 2.**

The associations of the PRS subgroups with LORA or YORA stratified by mLOY status were evaluated referring low PRS subgroup without mLOY in Set 2. ORs are indicated by dots, and 95%CIs are indicated by two-sided lines. PRS, polygenic risk score; mLOY, mosaic loss of chromosome Y; LORA, late-onset RA; YORA, young-onset RA.

**Supplementary Table S1. Demographic features of LORA and control subjects in each dataset**

|  | **Set 1: Discovery data** | | **Set 2: Validation data** | |
| --- | --- | --- | --- | --- |
|  | **LORA** | **Controls** | **LORA** | **Controls** |
| Number of subjects | 608 | 86,998 | 400 | 86,998 |
| Age (years), median (IQR) | 72.0 [68.0-76.3] | 65.0 [55.0-73.0] | 71.8 [67.0-75.5] | 65.0 [55.0-73.0] |
| Sex, n (%) |  |  |  |  |
| Male | 165 (27.1%) | 47,302 (54.4%) | 90 (22.5%) | 47,302 (54.4%) |
| Female | 443 (72.9%) | 39,696 (45.6%) | 310 (77.5%) | 39,696 (45.6%) |
| Smoking status, n (%) |  |  |  |  |
| Smokers | 210 (34.5%) | 43,194 (49.6%) | 149 (37.2%) | 43,205 (49.7%) |
| Never Smokers | 389 (64.0%) | 42,531 (48.9%) | 219 (54.8%) | 42,568 (48.9%) |
| No information | 9 (1.5%) | 1,273 (1.5%) | 32 (8.0%) | 1,225 (1.4%) |

**LORA; late-onset RA.**

**Supplementary Table S2. Demographic features of YORA and control subjects in each dataset**

|  | **Set 1: Discovery data** | | **Set 2: Validation data** | |
| --- | --- | --- | --- | --- |
|  | **YORA** | **Controls** | **YORA** | **Controls** |
| Number of subjects | 1,499 | 86,998 | 1,959 | 86,998 |
| Age (years), median (IQR) | 59.0 [51.5-65.0] | 65.0 [55.0-73.0] | 57.0 [49.0-63.5] | 65.0 [55.0-73.0] |
| Sex, n (%) |  |  |  |  |
| Male | 224 (14.9%) | 47,302 (54.4%) | 253 (12.9%) | 47,302 (54.4%) |
| Female | 1,275 (85.1%) | 39,696 (45.6%) | 1,706 (87.1%) | 39,696 (45.6%) |
| Smoking status, n (%) |  |  |  |  |
| Smokers | 482 (32.2%) | 43,194 (49.6%) | 621 (31.7%) | 43,205 (49.7%) |
| Never Smokers | 997 (66.5%) | 42,531 (48.9%) | 1,275 (65.1%) | 42,568 (48.9%) |
| No information | 20 (1.3%) | 1,273 (1.5%) | 63 (3.2%) | 1,225 (1.4%) |

**YORA; young-onset RA.**

**Supplementary Table S3. Demographic features comparing LORA and YORA in each dataset**

|  | **Set 1: Discovery data** | | **Set 2: Validation data** | |
| --- | --- | --- | --- | --- |
|  | **LORA** | **YORA** | **LORA** | **YORA** |
| Number of subjects | 608 | 1,499 | 400 | 1,959 |
| Age (years), median (IQR) | 72.0 [68.0-76.3] | 59.0 [51.5-65.0] | 71.8 [67.0-75.5] | 57.0 [49.0-63.5] |
| Age at onset (years), median (IQR) | 66.0 [62.0-71.0] | 44.0 [35.0-52.0] | 64.0 [61.0-68.0] | 45.0 [36.0-51.0] |
| Sex, n (%) |  |  |  |  |
| Male | 165 (27.1%) | 224 (14.9%) | 90 (22.5%) | 253 (12.9%) |
| Female | 443 (72.9%) | 1,275 (85.1%) | 310 (77.5%) | 1,706 (87.1%) |
| RF positive (%) | 346 (75.4%)  (n=459) | 936 (80.1%)  (n=1,168) | 328 (84.3%)  (n=389) | 1,688 (89.2%)  (n=1,892) |
| ACPA positive (%) | 368 (70.1%)  (n=525) | 1,126 (83.3%)  (n=1,351) | 314 (83.7%)  (n=375) | 1,645 (87.4%)  (n=1,882) |

**RF; rheumatoid factor, ACPA; anticitrullinated cyclic peptide antibody.**

**Supplementary Table S4. The associations between different types of mCAs and RA subsets (RA, LORA, and YORA) in each dataset corresponding to Figure S1**

|  | Set 1: Discovery data | | | | | Set 2: Validation data | | | | | | | Meta-analysis  (Set 1 and Set 2) | | |
| --- | --- | --- | --- | --- | --- | --- | --- | --- | --- | --- | --- | --- | --- | --- | --- |
|  |  | mCA carriers/  Participants (prevalence) | | OR  (95%CI) | P | mCA carriers/  Participants (prevalence) | | | OR  (95%CI) | | P | | OR  (95%CI) | | P |
|  |  | RA cases | non-RA controls |  |  | RA cases | non-RA controls |  | |  | |  | |  | |
| autosomal  mCAs | RA | 154/2,078  (7.41%) | 9,710/85,725  (11.33%) | 0.74  (0.62-0.89) | **0.00097** | 148/2,264  (6.54%) | 9,825/85,773  (11.5%) | 0.68  (0.57-0.82) | | **6.5×10^-5^** | | 0.71  (0.63-0.81) | | **2.7×10^-7^** | |
|  | LORA | 68/599  (11.4%) | 9,710/85,725  (11.33%) | 0.78  (0.59-1.02) | 0.066 | 43/368  (11.7%) | 9,825/85,773  (11.5%) | 0.76  (0.53-1.07) | | 0.12 | | 0.77  (0.62-0.95) | | **0.0158** | |
|  | YORA | 86/1,479  (5.81%) | 9,710/85,725  (11.33%) | 0.68  (0.53-0.85) | **0.0010** | 105/1,896  (5.54%) | 9,825/85,773  (11.5%) | 0.63  (0.51-0.79) | | **4.7×10^-5^** | | 0.65  (0.56-0.77) | | **1.9×10^-7^** | |
| mLOX | RA | 287/1,694  (16.9%) | 11,924/39,100  (30.5%) | 0.43  (0.37-0.49) | **9.7×10^-36^** | 460/1,928  (23.9%) | 12,018/39,098  (30.7%) | 0.78  (0.70-0.87) | | **1.8×10^-5^** | | 0.61  (0.55-0.66) | | **5.8×10^-30^** | |
|  | LORA | 118/437  (27.0%) | 11,924/39,100  (30.5%) | 0.53  (0.42-0.65) | **6.9×10^-9^** | 104/279  (37.3%) | 12,018/39,098  (30.7%) | 0.87  (0.67-1.11) | | 0.26 | | 0.65  (0.55-0.77) | | **3.1×10^-7^** | |
|  | YORA | 169/1,257  (13.4%) | 11,924/39,100  (30.5%) | 0.38  (0.32-0.45) | **3.7×10^-29^** | 356/1,649  (21.6%) | 12,018/39,098  (30.7%) | 0.76  (0.67-0.86) | | **2.2×10^-5^** | | 0.59  (0.53-0.65) | | **4.0×10^-24^** | |
| mLOY | RA | 99/384  (25.8%) | 11,179/46,625  (24.0%) | 1.10  (0.86-1.41) | 0.43 | 67/336  (19.9%) | 11,009/46,675  (23.6%) | 0.85  (0.64-1.14) | | 0.28 | | 0.99  (0.82-1.19) | | 0.91 | |
|  | LORA | 68/162  (42.0%) | 11,179/46,625  (24.0%) | 1.39  (1.00-1.93) | 0.050 | 37/89  (41.6%) | 11,009/46,675  (23.6%) | 1.26  (0.81-1.96) | | 0.31 | | 1.34  (1.03-1.75) | | 0.030 | |
|  | YORA | 31/222  (14.0%) | 11,179/46,625  (24.0%) | 0.73  (0.49-1.09) | 0.12 | 30/247  (12.1%) | 11,009/46,675  (23.6%) | 0.59  (0.39-0.88) | | **0.0093** | | 0.65  (0.49-0.87) | | **0.0034** | |

OR, odds ratio referring to non-RA controls; P value in bold indicate statistical significance based on Bonferroni correction (P<0.016); **LORA, late-onset RA; YORA, young-onset RA; mCAs, mosaic chromosomal alterations; mLOY, mosaic loss of chromosome Y; mLOX, mosaic loss of chromosome X.**

**Supplementary Table S5. Associations between the age of RA onset and mLOY among RA subjects**

|  | Dataset | OR (95%CI) | P |
| --- | --- | --- | --- |
| Onset age of RA | Set 1 | 1.02 (0.99-1.05) | 0.22 |
|  | Set 2 | 1.03 (1.00-1.07) | 0.063 |
|  | Meta-analysis  (Set 1 and Set 2) | 1.03 (1.00-1.05) | **0.031** |

P value in bold indicates statistical significance (P<0.05).

**Supplementary Table S6. The associations between cell fraction of mLOY >5% and RA subsets (RA, LORA, and YORA) in each dataset corresponding to Figure 4**

|  | Set 1: Discovery data | | | | | Set 2: Validation data | | | | Meta-analysis  (Set 1 and Set 2) | |
| --- | --- | --- | --- | --- | --- | --- | --- | --- | --- | --- | --- |
|  |  | mCA carriers/  Participants (prevalence) | | OR  (95%CI) | P | mCA carriers/  Participants (prevalence) | | OR  (95%CI) | P | OR  (95%CI) | P |
|  |  | RA cases | non-RA controls |  |  | RA cases | non-RA controls |  |  |  |  |
| mLOY | RA | 102/389 (26.2%) | 11,366/47,302 (24.0%) | 1.15  (0.90-1.46) | 0.27 | 68/343 (19.8%) | 11,172/47,302 (23.6%) | 0.88  (0.66-1.17) | 0.37 | 1.03  (0.85-1.23) | 0.79 |
| mLOY-CF >5% |  | 84/371  (22.6%) | 8,315/44,251  (18.8%) | 1.33  (1.02-1.73) | 0.040 | 52/327 (15.9%) | 8,197/44,327  (18.5%) | 0.94  (0.68-1.29) | 0.70 | 1.15  (0.94-1.41) | 0.18 |
| mLOY | LORA | 71/165  (43.0%) | 11,366/47,302 (24.0%) | 1.47  (1.07-2.04) | 0.020 | 38/90 (42.2%) | 11,172/47,302 (23.6%) | 1.36  (0.87-2.11) | 0.17 | 1.43  (1.10-1.86) | **0.0070** |
| mLOY-CF >5% |  | 62/156  (39.7%) | 8,315/44,251  (18.8%) | 1.69  (1.20-2.38) | **0.0027** | 33/85 (38.8%) | 8,197/44,327  (18.5%) | 1.51  (0.95-2.40) | 0.080 | 1.62  (1.23-2.14) | **0.00060** |
| mLOY | YORA | 31/224  (13.8%) | 11,366/47,302 (24.0%) | 0.73  (0.49-1.09) | 0.12 | 30/253 (11.9%) | 11,172/47,302 (23.6%) | 0.59  (0.39-0.88) | **0.0094** | 0.66  (0.49-0.87) | **0.0034** |
| mLOY-CF >5% |  | 22/215  (10.2%) | 8,315/44,251  (18.8%) | 0.75  (0.47-1.20) | 0.23 | 19/242 (7.85%) | 8,197/44,327  (18.5%) | 0.53  (0.33-0.87) | **0.010** | 0.64  (0.45-0.89) | **0.0090** |

OR, odds ratio referring to non-RA controls; P value in bold indicate statistical significance based on Bonferroni correction (P<0.016); **LORA, late-onset RA; YORA, young-onset RA; mLOY, mosaic loss of chromosome Y.**

**Supplementary Table S7. The associations between cell fraction of mLOY >5% and RA subsets (RA, LORA, and YORA) in each dataset corresponding to Figure S4**

|  | Set 1: Discovery data | | | | | Set 2: Validation data | | | | Meta-analysis  (Set 1 and Set 2) | |
| --- | --- | --- | --- | --- | --- | --- | --- | --- | --- | --- | --- |
|  |  | mCA carriers/  Participants (prevalence) | | OR  (95%CI) | P | mCA carriers/  Participants (prevalence) | | OR  (95%CI) | P | OR  (95%CI) | P |
|  |  | RA cases | non-RA controls |  |  | RA cases | non-RA controls |  |  |  |  |
| mLOY | RA | 99/384 (25.8%) | 11,179/46,625 (24.0%) | 1.10  (0.86-1.41) | 0.43 | 67/336 (19.9%) | 11,009/46,675 (23.6%) | 0.85  (0.64-1.14) | 0.28 | 0.99  (0.82-1.19) | 0.91 |
| mLOY-CF >5% |  | 82/367 (22.3%) | 8,193/43,639  (18.8%) | 1.27  (0.97-1.66) | 0.080 | 51/320 (15.9%) | 8,066/43,732 (18.4%) | 0.90  (0.66-1.25) | 0.54 | 1.11  (0.90-1.36) | 0.34 |
| mLOY | LORA | 68/162  (42.0%) | 11,179/46,625 (24.0%) | 1.39  (1.00-1.93) | 0.050 | 37/89 (41.6%) | 11,009/46,675 (23.6%) | 1.26  (0.81-1.96) | 0.31 | 1.34  (1.03-1.75) | 0.030 |
| mLOY-CF >5% |  | 60/154  (39.0%) | 8,193/43,639  (18.8%) | 1.59  (1.13-2.24) | **0.0084** | 32/84 (38.1%) | 8,066/43,732  (18.4%) | 1.38  (0.87-2.21) | 0.18 | 1.51  (1.15-2.00) | **0.0034** |
| mLOY | YORA | 31/222 (14.0%) | 11,179/46,625 (24.0%) | 0.73  (0.49-1.09) | 0.12 | 30/247 (12.1%) | 11,009/46,675 (23.6%) | 0.59  (0.39-0.88) | **0.0093** | 0.65  (0.49-0.87) | **0.0034** |
| mLOY-CF >5% |  | 22/213 (10.3%) | 8,193/43,639 (18.8%) | 0.75  (0.47-1.19) | 0.22 | 19/236 (8.1%) | 8,066/43,732 (18.4%) | 0.53  (0.32-0.87) | **0.011** | 0.63  (0.45-0.89) | **0.0085** |

OR, odds ratio referring to non-RA controls; P value in bold indicate statistical significance based on Bonferroni correction (P<0.016); **LORA, late-onset RA; YORA, young-onset RA; mLOY, mosaic loss of chromosome Y.**

**Supplementary Table S8. Male subjects used in the association analysis between mLOY and risk factors for RA with RA and the interaction analysis between mLOY and PRS.**

|  | Set 1: Discovery data | | Set 2: Validation data | |
| --- | --- | --- | --- | --- |
|  | RA cases | Controls | RA cases | Controls |
| Number of subjects | **389** | **45,369** | **341** | **45,412** |
| Smoking status, n (%) |  |  |  |  |
| Smokers | 323 (83.0%) | 33,686 (74.2%) | 287 (84.2%) | 33,701 (74.2%) |
| Never Smokers | 61 (15.7%) | 11,048 (24.4%) | 47 (13.8%) | 11,113 (24.5%) |
| No information | 5 (1.3%) | 635 (1.4%) | 7 (2.0%) | 598 (1.3%) |
| Types of RA, n (%) |  |  |  |  |
| LORA | 165 (42.4%) | - | 89 (26.1%) | - |
| YORA | 224 (57.6%) | - | 252 (73.9%) | - |

The bold numbers of samples remained after exclusion of related samples with PIHAT > 0.25; Of these samples, only samples whose smoking information were available were used for the association analysis between mLOY and risk factors for RA; **LORA, late-onset RA; YORA, young-onset RA.**

**Supplementary Table S9. Association between mLOY and RA-PRS in each dataset.**

|  | Dataset | OR (95%CI) | P |
| --- | --- | --- | --- |
| PRS | Set 1 | 1.01 (0.99-1.02) | 0.44 |
|  | Set 2 | 1.01 (0.99-1.02) | 0.33 |

PRS, polygenic risk score; mLOY, mosaic loss of chromosome Y.

**Supplementary Table S10. Associations of mLOY and RA-PRS with RA subsets in each dataset corresponding to Figure 5**

|  |  | Set 1 | | | Set 2 | | | Meta-analysis  (Set 1 and Set 2) | | | |
| --- | --- | --- | --- | --- | --- | --- | --- | --- | --- | --- | --- |
|  | RA subsets | beta | 95%CI | | beta | 95%CI | | beta | 95%CI | | P |
| PRS | RA | 0.52 | 0.46 | 0.58 | 0.58 | 0.52 | 0.65 | 0.55 | 0.51 | 0.59 | **3.3×10^-138^** |
|  | LORA | 0.43 | 0.34 | 0.52 | 0.45 | 0.33 | 0.58 | 0.44 | 0.37 | 0.51 | **1.7×10^-32^** |
|  | YORA | 0.58 | 0.51 | 0.66 | 0.63 | 0.56 | 0.70 | 0.61 | 0.55 | 0.66 | **3.0×10^-111^** |
| mLOY | RA | 0.11 | -0.14 | 0.35 | -0.11 | -0.40 | 0.17 | 0.01 | -0.17 | 0.20 | 0.90 |
|  | LORA | 0.33 | 0.003 | 0.66 | 0.30 | -0.14 | 0.75 | 0.32 | 0.06 | 0.58 | **0.017** |
|  | YORA | -0.32 | -0.72 | 0.082 | -0.50 | -0.90 | -0.10 | -0.41 | -0.69 | -0.13 | **0.0047** |

P-values in bold indicate statistical significance (P<0.05); PRS, polygenic risk score; **LORA, late-onset RA; YORA, young-onset RA; mLOY, mosaic loss of chromosome Y.**

**Supplementary Table S11. Associations of mLOY and RA-PRS with LORA in each dataset corresponding to Figure S5**

|  | Set 1 | | | Set 2 | | | Combined dataset  (Set 1 and Set 2) | | | |
| --- | --- | --- | --- | --- | --- | --- | --- | --- | --- | --- |
|  | beta | 95%CI | | beta | 95%CI | | beta | 95%CI | | P |
| PRS | -0.15 | -0.32 | 0.01 | -0.17 | -0.37 | 0.03 | -0.16 | -0.29 | -0.03 | **0.014** |
| mLOY | 0.11 | -0.54 | 0.76 | 0.91 | 0.17 | 1.65 | 0.46 | -0.03 | 0.95 | 0.067 |

The p-value in bold indicates statistical significance (P<0.05). **PRS, polygenic risk score; mLOY, mosaic loss of chromosome Y.**

**Supplementary Table S12. Results of the one-way ANOVA to evaluate the goodness of fit between the two models**

|  | Residual deviance | Deviance | P |
| --- | --- | --- | --- |
| $model1$ | 3356.9 |  |  |
| $model2$ | 3348.4 | 8.48 | **0.0036** |

$model1: LORA～ mLOY+PRS group$

$$model2: LORA～ mLOY+PRS group+PRS group\times mLOYCF>5\%$$

The p-value in bold indicates statistical significance (P<0.05).
